# COVID-19 related messaging, beliefs, information sources, and mitigation behaviors in Virginia: A cross-sectional survey in the summer of 2020

**DOI:** 10.1101/2021.08.18.21262217

**Authors:** Rachel A. Silverman, Danielle Short, Sophie Wenzel, Mary Ann Friesen, Natalie E. Cook

## Abstract

Conflicting messages and misinformation related to the coronavirus (COVID-19) pandemic (SARS-CoV-2) have hindered mitigation efforts. To gain insight and inform effective evidence-based public health messaging, we distributed an online cross-sectional survey from May to July, 2020. Among 3,488 respondents, systematic differences were observed in information sources that people trust, events that impacted beliefs and behaviors, and how behaviors changed by socio-demographics, political identity, and geography within Virginia. Characteristics significantly associated (p<0.05) with not wearing a mask in public included identifying as non-Hispanic white, men, Republican, younger age, lower income, not trusting national science and health organizations, believing a non-evidence-based messages, and Southwest Virginia in logistic regression. Similar, lesser in magnitude correlations, were observed for distancing in public. This study can assist decision makers and the public to improve and effectively target public health messaging related to the ongoing COVID-19 pandemic and future public health challenges in Virginia and similar jurisdictions.

## Introduction

The early days of the coronavirus (COVID-19) pandemic (SARS-CoV-2) were characterized by conflicting messaging from nearly all levels of national and international mass media and government [1-4]. As public health and healthcare professionals attempted to quell the growing panic with science-driven narratives, conspiracy theories and misinformation continued to spread through social media platforms such as Facebook, Weibo, and Twitter, often undermining or contradicting the life-saving messages that scientists were trying to communicate [5, 6]. This issue was further compounded by long-standing health, socioeconomic, and racist inequities as well as sharp decreases in funding to state and federal health agencies in the United States [5]. Throughout the pandemic, access to and acceptance of evidence-based messaging to prevent and respond to outbreaks of coronavirus disease (COVID-19) have been inconsistent across populations [7]. Black, Hispanic, and Indigenous populations have been historically excluded from the United States’ public health and medical institutions, often suffering disproportionately from many diseases and public health challenges [8, 9]. When combined with the knowledge that ethnic minority, low-income, low-education, and elderly populations are overrepresented in COVID-19 related morbidity and mortality numbers, public health officials will need to effectively reach out to and target those particular groups[10-12].

Several surveys have evaluated the awareness and concern that members of the public have experienced towards COVID-19 and local, state, and national government responses [7, 13]. Results showed that the majority of the general population wants to hear from public health and medical officials, and are likely to trust professional sources that have self-protective and pro-social messages that focus on positive ways to protect themselves and their loved ones [14, 15]. This includes demographic groups that are considered high-risk for COVID-19, like the elderly and low-income individuals from minority groups [16-18].

As vaccine coverage increases at different rates globally, the public health response to COVID-19 continues to necessitate coordination at all levels of government to ensure accessible and accurate testing, contact tracing, quarantine and isolation, treatment, and mitigation measures like social or physical distancing and mask wearing [19]. It is important that trust in public health information be maintained for these strategies to continue to be implemented effectively. Studies found that trust in public health officials and the information they provided allowed for successful messaging campaigns with past disease outbreaks, ranging from food safety incidents to worldwide polio vaccination campaigns [17]. As shown in past responses to foodborne disease outbreaks, demonstrating that public health measures and preventative strategies are in the best interests of the community overall is crucial to building and maintaining public trust that is essential to effective public health guidance [20]. High-risk populations may respond best when targeted with official messages that are consistent, credible, proactive, and also a mixture of self-focused and prosocial [14].

Throughout the COVID-19 pandemic in the United States, social distancing, school closures, lockdowns, and targeted public health messaging have been sporadic and inconsistent. Many people obtain information from social media that can conflict with the messages from public health officials [6]. In response, Facebook, Twitter, and online newspapers are now actively monitoring their own sites for COVID-19 disinformation that could mislead people into believing potentially dangerous rumors, stigmas, and conspiracy theories [6, 21]. The rapid development and rollout of COVID-19 vaccines have also been subject to misinformation on social media platforms, with peer-networks exchanging large quantities of anti-vaccination posts that focus on adverse side-effects, misleading medical content, and unsubstantiated rumors [22]. Similar methods have been used to undermine prior vaccine campaigns, and developing effective messaging to counter such disinformation will likely prove to be an important challenge for public health officials [23, 24].

Like many other large states, Virginia has had notable regional differences in case trends over time, with the more densely populated northern and central regions experiencing large case increases during the pandemic’s initial wave in the spring of 2020, while the coastal eastern region and the more rural southwestern region experiencing their first large case increases mid-summer [25]. Some Virginia college towns, such as Charlottesville, Blacksburg, and Harrisonburg, saw increases in local case counts when students returned in the late summer and mid-winter of 2020-21, showing that the movement of large groups of people can greatly affect community spread in less densely populated areas [26-28]. Given the continued need for effective evidence-based public health messaging, officials, researchers, and the public can benefit from exploring how people receive information they believe and trust, and how their beliefs influence their behaviors. To gain better insight, we conducted a cross-sectional survey during the summer of 2020 to examine COVID-19 related messaging, beliefs, information sources, and mitigation behaviors among adults in Virginia.

## Materials and Methods

We surveyed a convenience sample of Virginia residents by distributing a link to complete the survey online through our professional and personal email listservs, on Facebook, and on flyers in select locations. Eligibility criteria included being 18 years of age or older and residing in Virginia. Participants provided electronic informed consent prior to beginning the survey. The survey collected socio-demographic information, including gender, age, race, ethnicity, level of education, income, employment status, occupation, changes in employment due to the pandemic, political affiliation, sexual orientation, and zip code. Participants were asked about their perceptions of COVID-19, risk mitigation behaviors, messages and events they felt influenced their beliefs and behaviors, and where they obtained information that they trust. The full survey is available in the supplement. The survey was developed and administered using Qualtrics. Participants were able to save and continue the survey and the *Prevent Ballot Box Stuffing* option was selected in the survey settings to limit people from completing the survey more than once. Responses were completely anonymous.

For this analysis, we conducted exploratory analyses by calculating descriptive statistics of survey responses and investigated correlations between information sources, perceptions, beliefs, and risk mitigating behaviors related to the COVID-19 pandemic using Pearson’s Chi^2^ test (alpha of 0.05). We also investigated correlates of the fundamental risk mitigating behaviors mask wearing and social/physical distancing in unadjusted and adjusted analyses using logistic regression with robust variance estimates (alpha of 0.05). We adjusted for race, political identity, gender, age group, income, reporting national science and health organizations as an information source, believing in alternative messages, and living in southwest Virginia to identify the independent effects of these characteristics on risk mitigation behaviors. These variables were selected a priori as known correlates of COVID-19 beliefs and incidence [10, 18, 29-36]. Data from surveys were excluded only if none of the questions beyond eligibility were answered.

All analyses were conducted using Stata/SE 16.1 and Microsoft Excel. This work was conducted by the Community and Collaborative subgroup of the integrated Translational Health Research Institute of Virginia (iTHRIV), a collaboration between Virginia Tech, University of Virginia, Inova, and Carilion Clinic. This study was approved by the Virginia Tech institutional Review Board (IRB number: 20-353) and the Inova Institutional Review Board (IRB number: U20 05-4056), prior to initiation of study activities at the respective sites.

## Results

### Respondent characteristics

The survey was open from May 19th to July 19th, 2020. Of the 3,694 individuals who started the survey, 3,678 (99.6%) self-reported as eligible and of these 190 (5%) did not answer any survey questions and were excluded. Of the remaining 3,488 respondents 3,367 (97%) fully completed the survey. Of the 3,488 included in this analysis, 70% completed the survey in May, 21% in June, and 9% in July of 2020. Participants were represented throughout Virginia (See Fig 1), with the largest numbers of respondents residing in Montgomery County (home of Virginia Tech), Loudoun and Fairfax Counties (near Washington DC, home of Inova), and Wise County (home of UVA Wise), reflecting sites where survey recruitment began.

**Figure 1.**
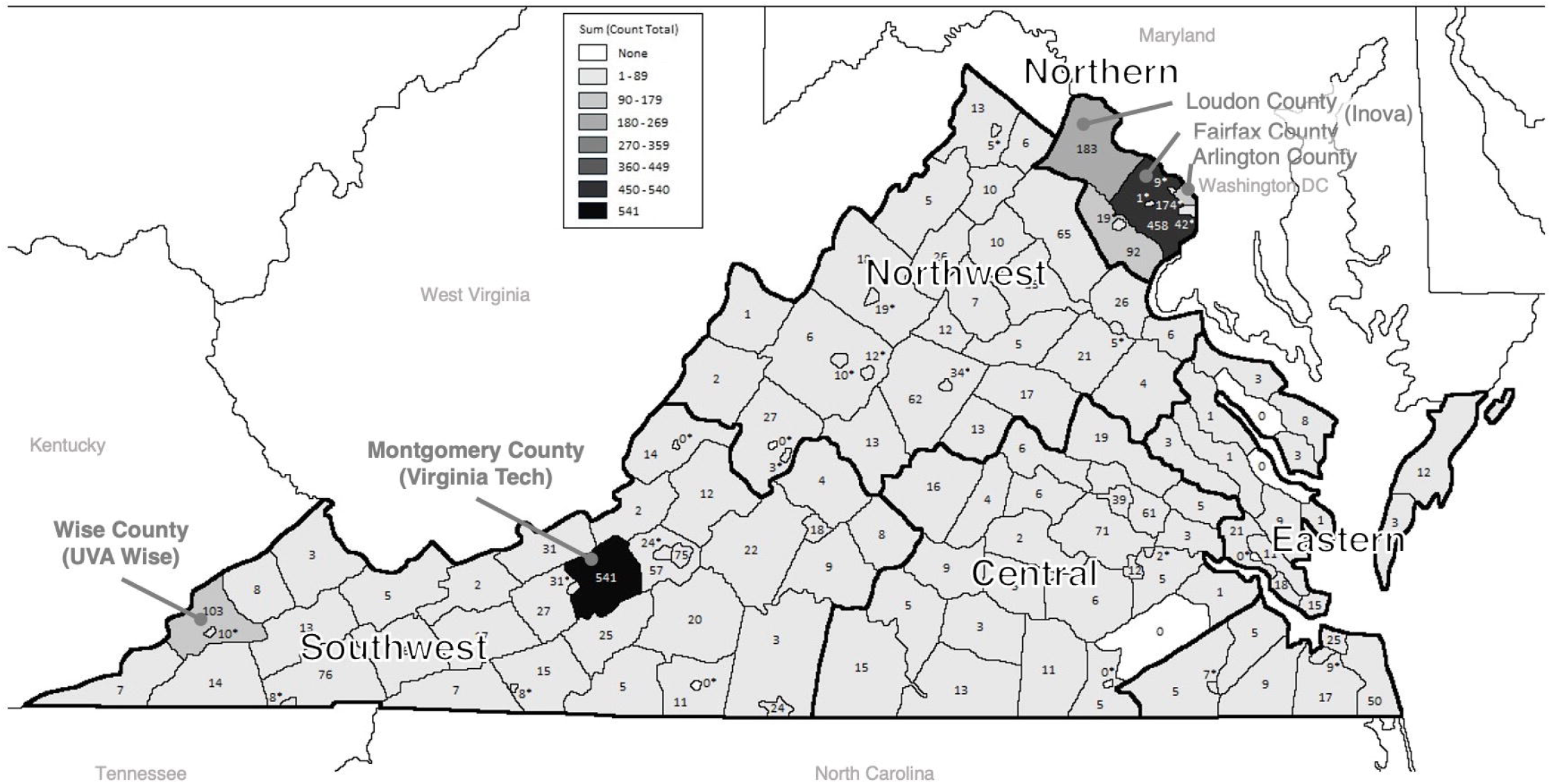
Map of Survey Respondents by County in Virginia (N=3,307). Number of respondents for each county in Virginia.

Sociodemographic characteristics are presented in Table 1. Briefly, 78% were women, 3% were Hispanic, 83% were non-Hispanic White, 5% were Black, 3% were Asian, and 87% identified as heterosexual or straight. Six percent were 18-24 years old, 6% were 25-29, 16% were 30-39, 19% were 40-49, 20% were 50-59, 20% were 60-69, and 9% were 70 or older. Most (94%) had completed at least some college or other post-high school education or training. Forty-three percent reported annual household income at least $100,000, 11% between $80,000 and $100,000, 13% between $60,000 and $80,000, 12% between $40,000 and $60,000, and 11% less than $40,000. Forty-six percent of respondents identified as Democrat, 13% as Republican, 22% as independent, and 13% said other or no-preference. Employment status was not mutually exclusive and 58% had full-time employment, 12% had part-time employment, 16% were retired, 5% were students, and 4% were unemployed. Eighteen percent of respondents reported a loss of or reduced employment or income, while 70% reported no change in their employment status as a result of COVID-19.

**Table 1.**
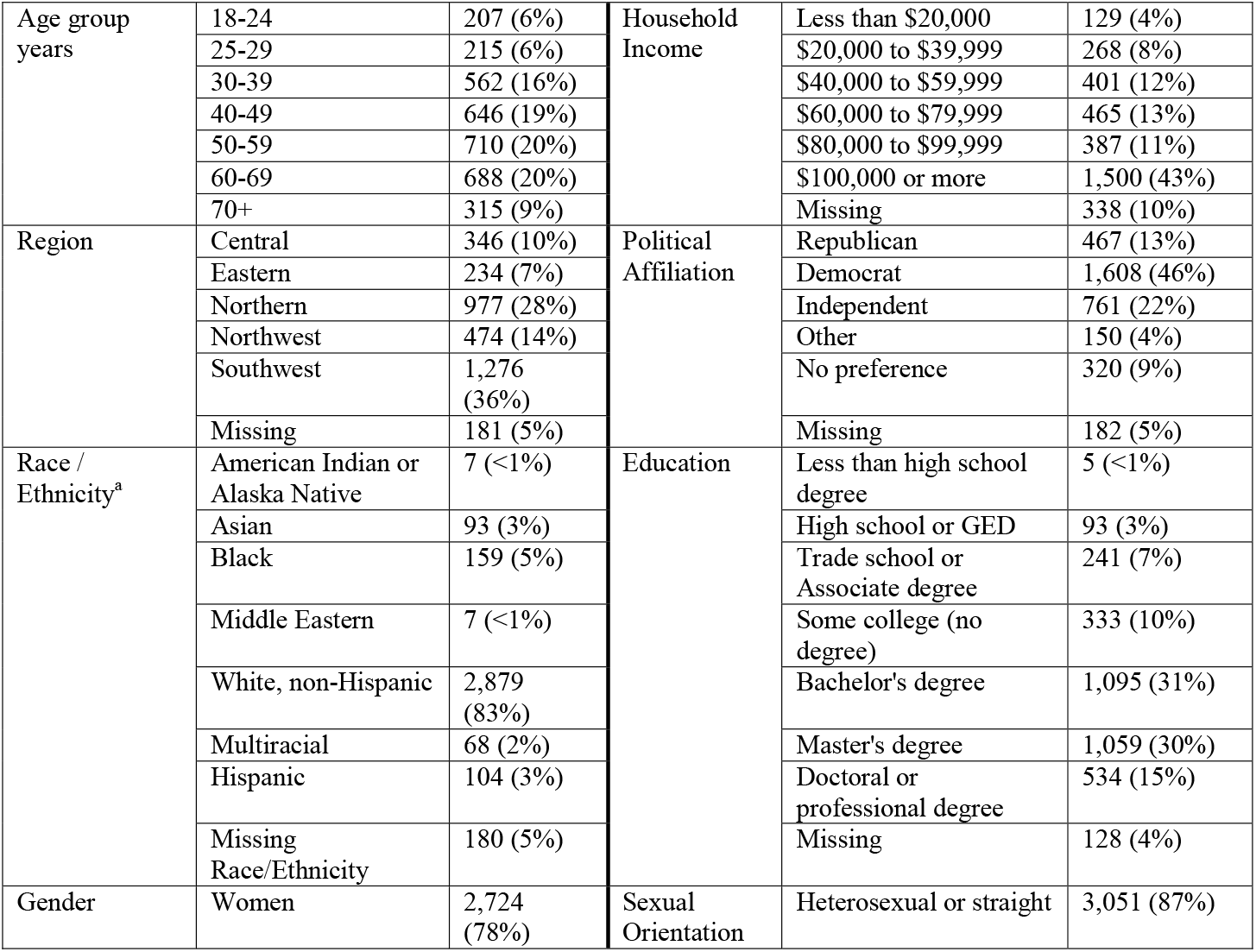
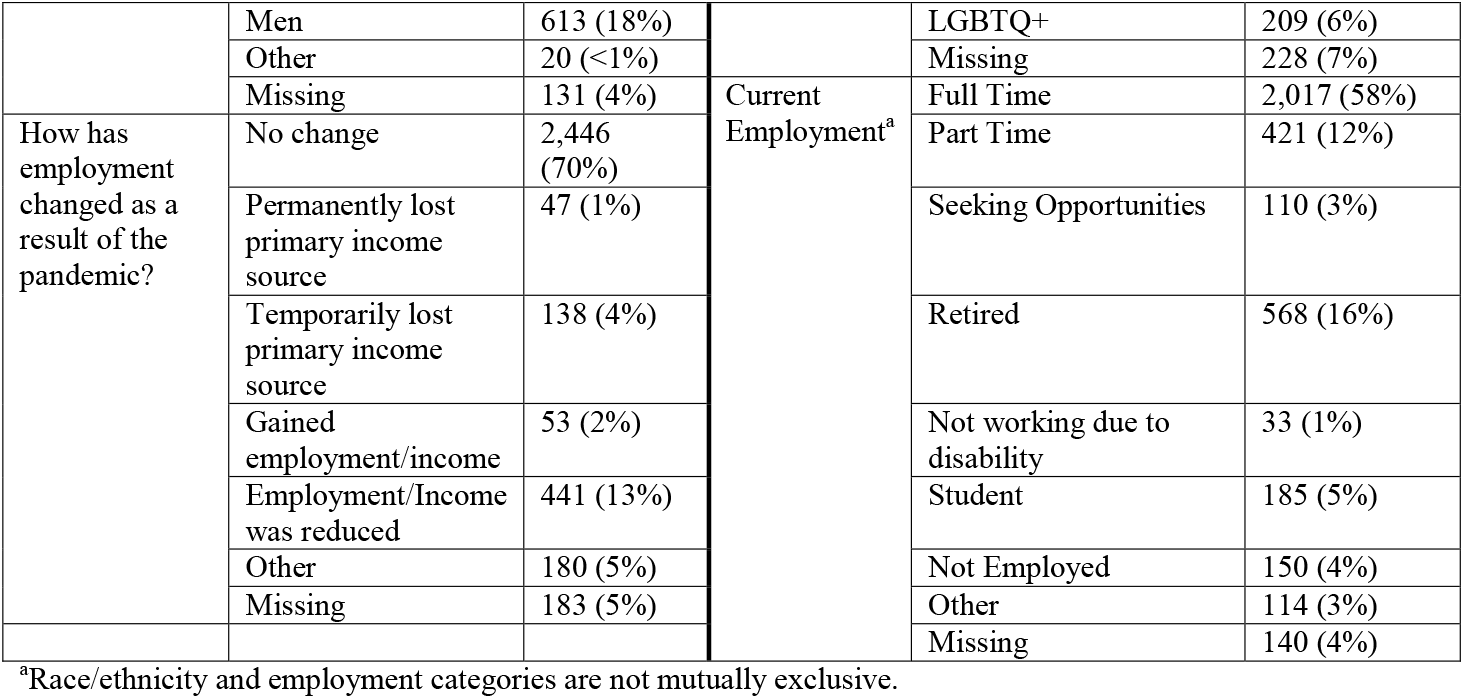
Characteristics of survey respondents (N = 3,488).

### Trusted information sources

All but 16 respondents (0.5%) answered the question: “Where do you get information that you trust about coronavirus/COVID-19?” Of these (n=3,472), most reported national science and health organizations (85%) as a trusted source for COVID-19 information and over 50% of respondents reported state/local health departments (75%), healthcare professionals (74%), and online news sources (55%) as a trusted source. Information sources reported by less than half of respondents included family and friends (26%), faith leader (4%), local TV news (34%), national TV news (49%), printed newspaper (20%), radio (20%), social media (22%), local government leaders (46%), and federal government leaders (22%). Only 2% of these respondents reported not following any COVID-19 updates (see Fig 2a).

**Figure 2.**
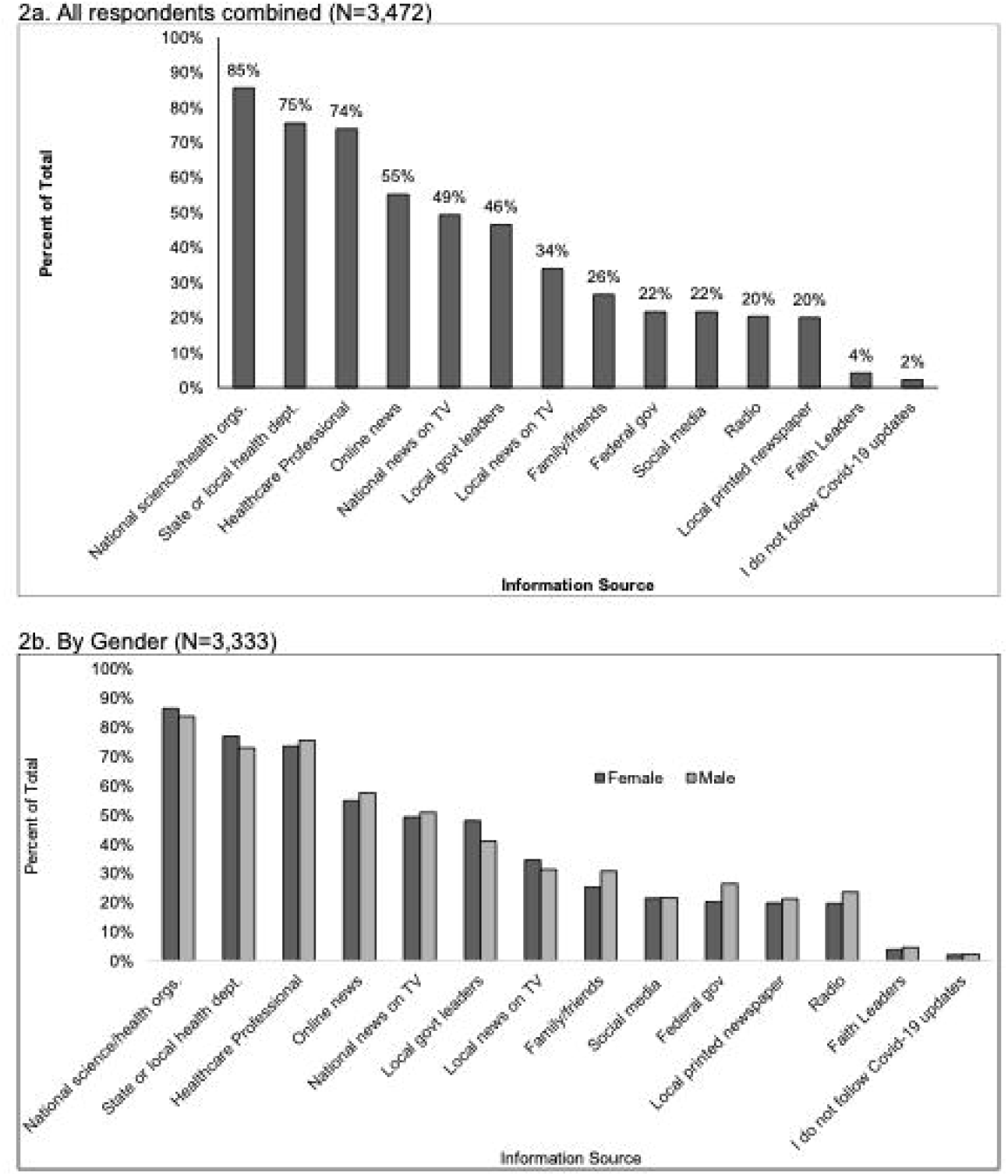
Where respondents received information that they trust about COVID-19, by participant characteristics. Survey responses to the question: “Where do you get information that you trust about coronavirus/COVID-19? (Check all that apply)” for all respondents (2a), and by gender (2b), age-group (2c), race/ethnicity (2d), political identity (2e), education level (2f), income level (2g), and Virginia region (2h).

More women than men received information they trusted from local government leaders (48% vs. 41%), and state or local health departments (77% vs. 73%) and more men than women received information they trusted from family/friends (31% vs. 25%), and federal government leaders (26% vs. 20%), all of which were statistically significantly difference (p<0.05) (see Fig 2b). Young adults age 18-24 were more likely than those of all older ages combined to receive trusted information from family and friends (39% vs. 25%) and social media (31% vs. 21%) and less likely from printed news (6% vs. 21%), radio (10% vs. 21%), and local government leaders (36% vs. 47%) (see Fig 2c). Slightly less non-Hispanic White and Asian respondents received trusted information from faith leaders (4% and 3%, respectively) than other races including 9% of Black and 10% of multiracial respondents. Non-White were also more likely than White respondents to receive information from local TV news (42% vs. 33%), and social media (27% vs. 21%) (see Fig 2d). White respondents were more likely than non-White respondents to receive information from printed local newspaper (21% vs. 14%), national science and health organizations (87% vs. 82%), or state and local health departments (77% vs. 71%) (see Fig 2d). More Democrats than Republicans received information they trusted from national science and health organizations (93% vs. 72%, respectively), State or local health department (83% vs. 62%), online news (65% vs. 41%), national TV news (55% vs. 43%), local government leaders (54% vs. 46%), printed newspaper (45% vs. 13%) and radio (26% vs. 14%) (see Fig 2e). More Republicans than Democrats received information they trusted from federal government leaders (46% vs. 13%), faith leaders (6% vs. 3%), or did not follow coronavirus/COVID-19 updates (5% vs. 0.3%). Similar proportions by political identity received information from local TV news, family/friends, healthcare-professionals, and social media. Other differences presented in Fig 2 were not statistically significant (p≥0.05).

More of those with than without a college degree received information from local printed newspapers (22% vs. 13%), radio (22% vs. 15%), online news (58% vs. 44%), local government leaders (47% vs. 43%), national science and health organizations (89% vs. 74%), and State or local health departments (79% vs. 64%) (see Fig 2f). More of those without than with a college degree received information from local faith leaders (6% vs. 4%), local TV news (38% vs. 33%), and federal government leaders (26% vs. 20%). More higher- than middle- and lower-income individuals received information from local printed newspapers (23%, 18%, 15%), online news (58%, 55%, 51%), local government leaders (49%, 43%, 45%), national science and health organizations (89%, 84%, 83%), and State or local health departments (80%, 77%, 70%) (see Fig 2g). Differences in information sources were also observed across regions (see Fig 2h).

### Perceptions & Beliefs Related to COVID-19

Twenty-three percent of respondents reported being “very worried” about catching COVID-19 and 34% were “very worried” about experiencing severe disease or complications if they were to catch COVID-19. Most respondents considered COVID-19 to be very serious (82%) or somewhat serious (13%). Statistically significant differences (p<0.05) were found between women vs. men (85% vs. 77%), Democrats vs. Republicans vs. others (95% vs. 61% vs. 78%), those 60 or more years old vs. those under 60 years old (91% vs. 81%), LGTBQ+ vs. heterosexual (89% vs. 83%), and higher-income vs. lower-income (86% reporting $100,000 vs. 78% making less than $20,000), and those with vs. without a college degree (86% vs. 75%) considered COVID-19 to be very serious. Other demographic characteristics were not statistically significantly different in terms of perceived seriousness of COVID-19. Differences by race and ethnicity were also observed with 91% of Asian, 87% of Black and Hispanic, 84% of multiracial and White, non-Hispanic respondents reporting they believed COVID-19 to be very serious.

Among those who answered the question: “Which if any of the following impacted your belief that COVID-19 was serious” (n=3,371), more than half selected hearing about COVID-19 in other countries (77%) or other states (73%), public-school closings (69%), the governor recommending a stay-at-home order (51%), mandating a stay-at-home order (65%), and declaring a state of emergency (68%), the CDC recommending that everyone wear a face mask in public (67%), restaurant dining rooms shutting down, (58%), and sporting events being canceled or postponed (52%) (see Fig 3a). Less than half selected becoming sick (3%) or knowing someone who became sick (30%), being high-risk or living with some-one who is high-risk for severe disease (40%), starting to work from home (35%), being laid off or losing their job (5%), when stores began limiting purchases of essential items (42%), when religious services were moved online (40%), and when a public figure tested positive (17%). Two percent of those who responded to this question said they did not think COVID-19 is serious, and 11% selected some other reason not listed in the survey.

**Figure 3.**
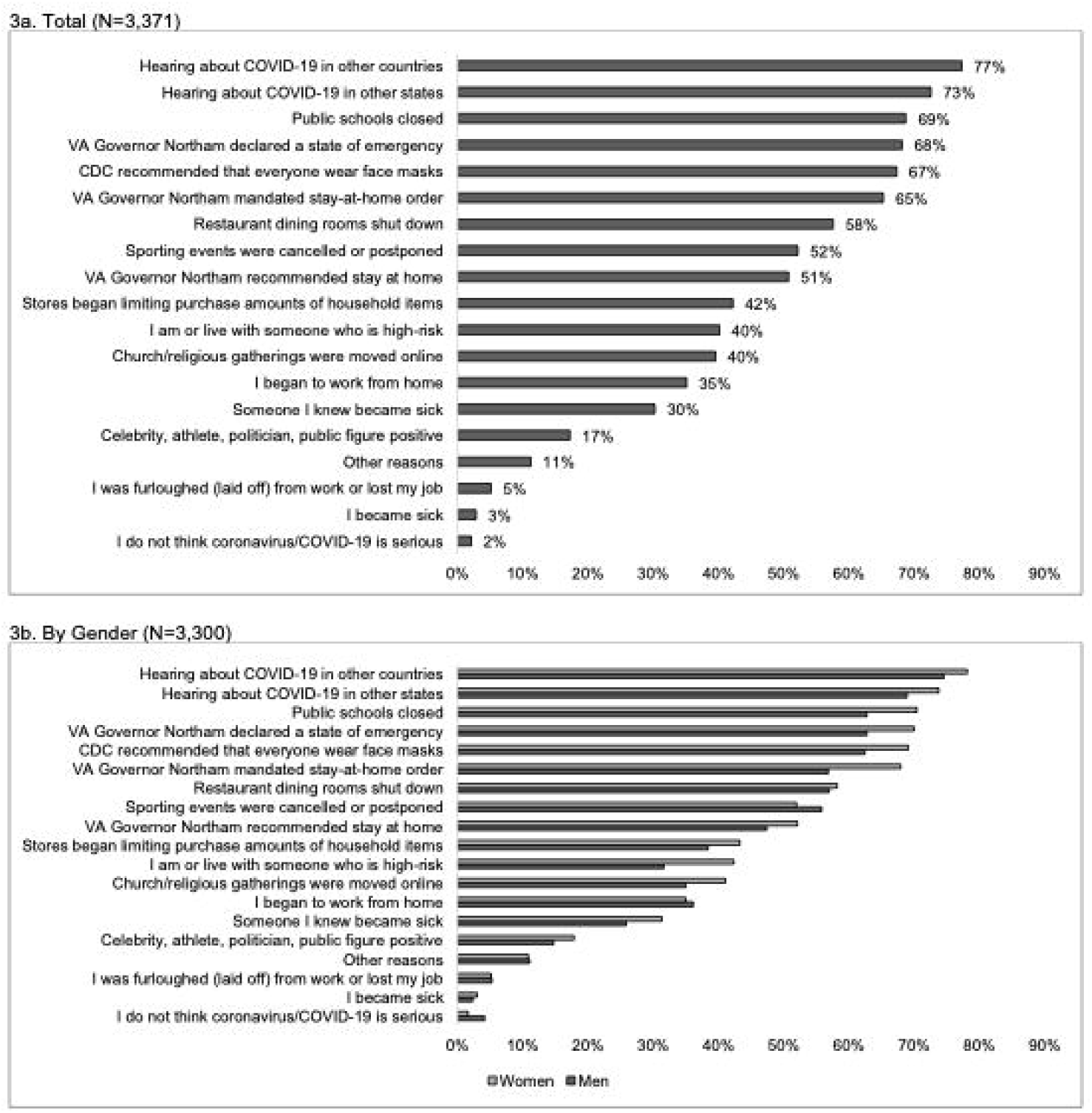
Events impacting belief that COVID-19 was serious, by participant characteristics. Survey responses to the question: “Which (if any) of the following have affected whether or not you think the coronavirus/COVID-19 is serious? (Check all that apply)” for all respondents (3a), and by gender (3b), age-group (3c), race/ethnicity (3d), political identity (3e), education level (3f), income level (3g), and Virginia region (3h).

Among those who answered the question: “Which if any of the following impacted your belief that COVID-19 was serious,” women were statistically significantly (p<0.05) more likely than men to state someone they knew became sick (31% vs. 26), being or living with someone high-risk (42% vs. 32%), public schools closing (71% vs. 63%), the governor declaring a state of emergency (70% vs. 63%), the CDC recommending face masks (69% vs. 63%), hearing about it in other countries (78% vs. 75%) and states (74% vs. 69%), stores limiting purchases (43% vs. 38%), religious services gong online (41% vs. 35%), and the governor recommending a stay at home order (52% vs. 47%) and mandating stay at home order (68% vs 57%) (see Fig 3b). Young adults were more likely than all other age-groups combined to select restaurant dining rooms (66% vs. 57%), being furloughed (laid off) or losing their job (16% vs. 4%), stores limiting purchases, (52% vs. 42%), and a public figure testing positive (26% vs. 17%) and less likely to select hearing about COVID-19 in other countries (72% vs. 78%), the governor *recommending* a stay-at-home order (41% vs. 52%), though this is not true for when the governor *mandated* the stay-at-home order (68% vs 66%) compared to other age groups (see Fig 3c). Non-white people were more likely than white people to select personally becoming sick (6% vs. 2%) or someone they knew becoming sick (45% vs. 28%), stores limiting purchases (52% vs. 41%), religious services moving online (47% vs. 39%), and a public figure testing positive (23% vs. 17%) (see Fig 3d). Democrats were more likely than Republicans to select knowing someone who was sick (32% vs. 24%), public schools closing (74% vs. 62%), restaurants closing (61% vs. 53%), the governor declaring a state of emergency (80% vs. 45%), starting to work from home (40% vs. 29%), the CDC recommending face masks (76% vs. 53%), hearing about COVID-19 in other countries (86% vs. 59%) and states (81% vs. 58%), stores limiting purchases (46% vs. 38%), sporting events being canceled or postponed (56% vs. 48%) and a public figure testing positive (20% vs. 13%) (see Fig 3e). More Republicans than Democrats selected religious services moving online (45% vs. 37%). More of those with than without a college degree reported their believe was impacted by the governor declaring a state of emergency (70% vs. 63%), beginning to work from home (38% vs. 24%), the CDC recommending face masks (69% vs. 63%), hearing about COVID-19 in other countries (81% vs. 65%) or states (75% vs. 65%), and sporting events being cancelled (53% vs. 49%) (see Fig 3f). More of those without than with a college degree reported their believe was impacted by becoming sick (5% vs. 3%), being or living with someone who is high-risk (47% vs. 39%), being furloughed (laid off) from work (9% vs. 4%), stores limiting purchases (46% vs. 41%) and religious services moving online (46% vs. 38%).

Fewer higher-income (>$100,000) than middle-income ($60,000-$99,999) and lower-income (<$60,0000) reported their belief was impacted by being or living with someone who is high-risk (37%, 41%, 44%), being furloughed (laid off) from work or losing their job (4%, 5%, 9%), stores limiting purchases of essential items (37%, 46%, 48%), religious gatherings being moved online (36%, 42%, 41%), a celebrity getting testing positive for COVID-19 (16%, 16%, 22%), and/or the governor recommending a stay at home order (49%, 55%, 52%). More higher-than middle- and lower-income reported beginning to work from home (39%, 35%, 32%), and hearing about COVID-19 in other countries (80%, 77%, 75%) (see Fig 3g). Differences in what impacted the beliefs that COVID-19 was serious were also observed across regions (see Fig 3h), with statistically significant differences (p<0.05) observed for becoming sick, knowing someone who became sick, being or living with someone at high risk, beginning to work from home, and sporting events being cancelled or postponed.

### Evidence based and alternative messages related to COVID-19

Over 80% of respondents reported that one or more of the following evidence-based messages impacted their beliefs and/or behaviors: “the coronavirus is highly contagious;” “stay home stay safe;” “stay home, save lives, slow the spread;” “practice social distancing;” “don’t touch your face;” and “wash your hands for at least 20 seconds” (see Fig 4). Twelve percent of respondents reported believing in one or more of the following alternative messages: COVID-19 “was developed as a bioweapon” (6%), “was developed to lower social security payments to seniors” (1%), “is a sign of the apocalypse/end times” (3%), “is a hoax” (1%), “can be treated with natural remedies” (3%), “was developed for population control” (3%), and/or “was developed to increase sales of cleaning supplies” (4%).

**Figure 4.**
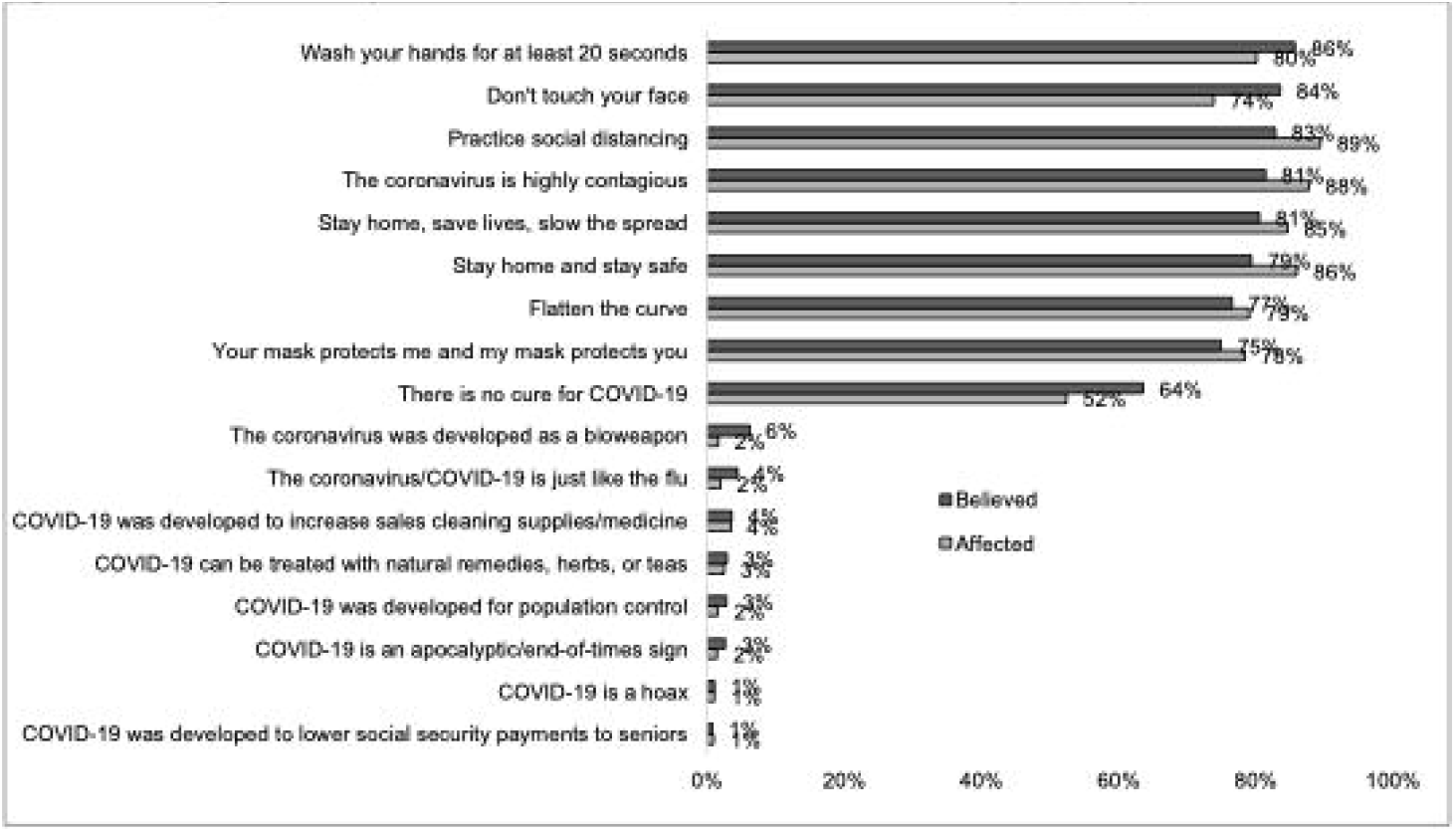
Messages that respondents believe and/or affected their behaviors (N=3,445). Survey responses to the question: “The following messages are related to the coronavirus/COVID-19 (not all are true). Please check all that apply if you have heard, believe, and/or changed your behavior based on each message” for all respondents.

### Correlations between alternative messages, demographics, and information sources

The same proportion (12%) of men and women (see Fig 5a), more young adults vs. older ages combined (22% vs. 12%), with the lowest among those 70 years old and greater (5%) (see Fig 5b), less of those who identified as non-Hispanic White (10%) compared to other races/ethnicities including 37% of Black, 22% of multiracial, 21% of Hispanic, and 15% of Asian (see Fig 5c), more Republicans (24%) than Democrats (7%) and others (15%) (see Fig 5d) believed in one of more alternative messages. The percent of those who believed in one of more alternative messages decreased with increasing education (29% of high-school degree or less to 6% of those with a doctoral degree) (see Fig 5e), and with increasing income level (9%, 13%, 17%) (see Fig 5f). More of those in Central Virginia than other regions (20% vs. <14%) (see Fig 5g) believed in one or more alternative messages.

**Figure 5.**
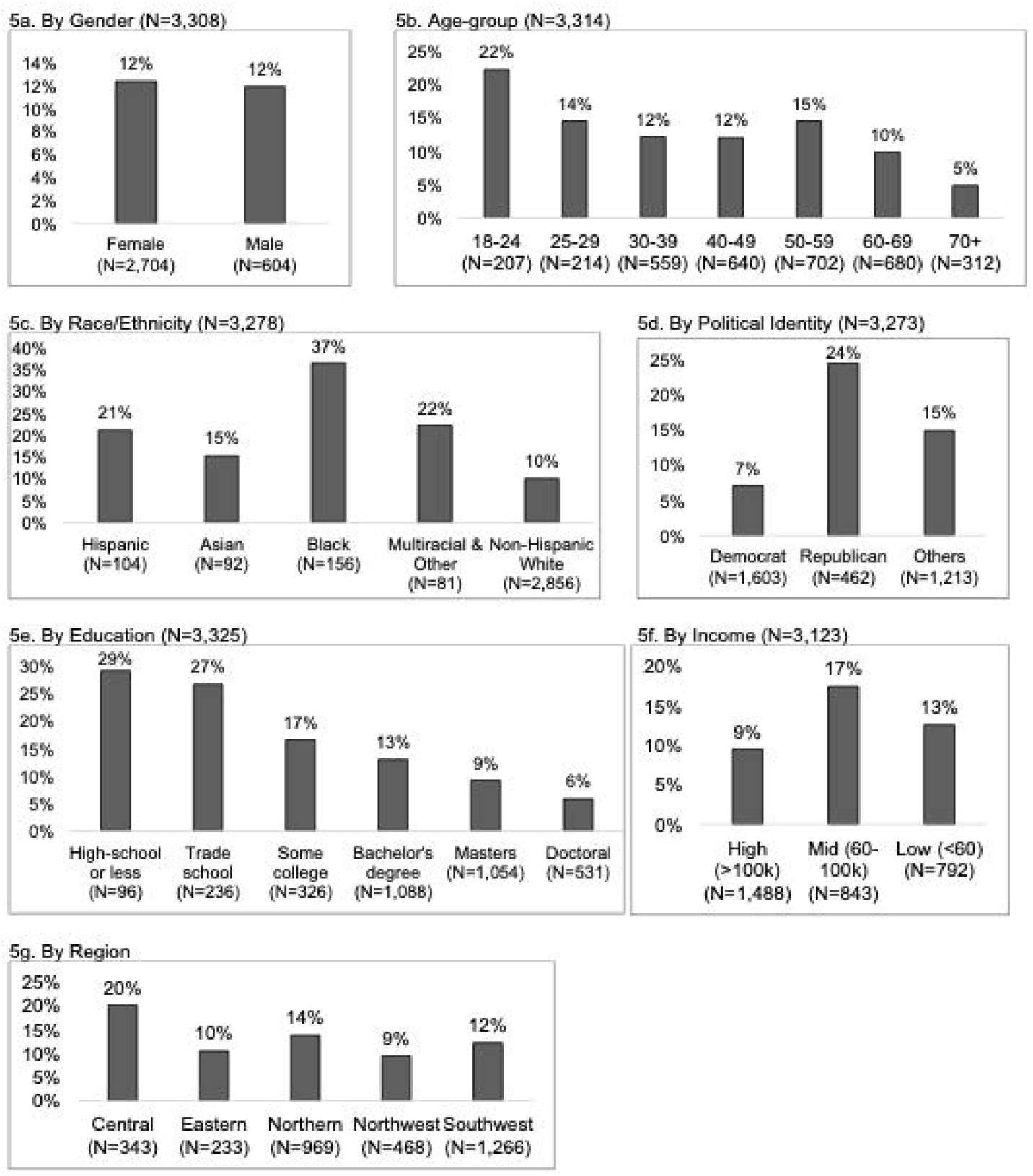
Percent that believed one or more alternative messages*, by participant characteristics. Percent of respondents who selected they believed in one or more alternative message when answering the question: “The following messages are related to the coronavirus/COVID-19 (not all are true). Please check all that apply if you have heard, believe, and/or changed your behavior based on each message” by gender (5a), age-group (5b), race/ethnicity (5c), political identity (5d), education level (5e), income level (5f), and Virginia region (5g). Alternative messages response options include: COVID-19 “was developed as a bioweapon,” “was developed to lower social security payments to seniors,” “is a sign of the apocalypse/end times,” “is a hoax,” “can be treated with natural remedies,” “was developed for population control,” and “was developed to increase sales of cleaning supplies.”

Those who believed in an alternative message were statistically significantly more likely than those who did not to receive trusted information from family and friends (32% vs. 26%), a faith leader (8% vs. 3%), local TV news (38% vs. 33%), social media (29% vs 21%), federal government leaders (33% vs. 20%), or report not following COVID-19 updates (see Fig 5h). Those who did not believe any alternative messages were more likely than those who did to receive trusted information from a healthcare professional (74% vs. 69%), local newspaper (21% vs. 13%), radio (21% vs. 15%), online news (57% vs, 45%), local government leaders (48% vs. 38%), national science and health organizations (88% vs. 67%), and state or local health departments (78% vs. 59%).

### Risk mitigation behavior changes

Ninety-eight percent of respondents completed the questions about changes in behaviors and 98% of those reported changing their behavior in some way in response to the pandemic (see Fig 6a). More than half of respondents reported one or more of the following behavior changes: practicing social/physical distancing (95%), wearing a mask when in public (90%), washing hands more often (90%), shopping for groceries and other essentials less often (86%), washing hands for 20 seconds (86%), being more careful not to touch their face in public (82%) and/or with unwashed hands (80%), using hand sanitizer more often (79%), avoiding public spaces (73%), cleaning frequently touched surfaces (68%), stocking up on supplies (62%), and started working from home (52%).

**Figure 6.**
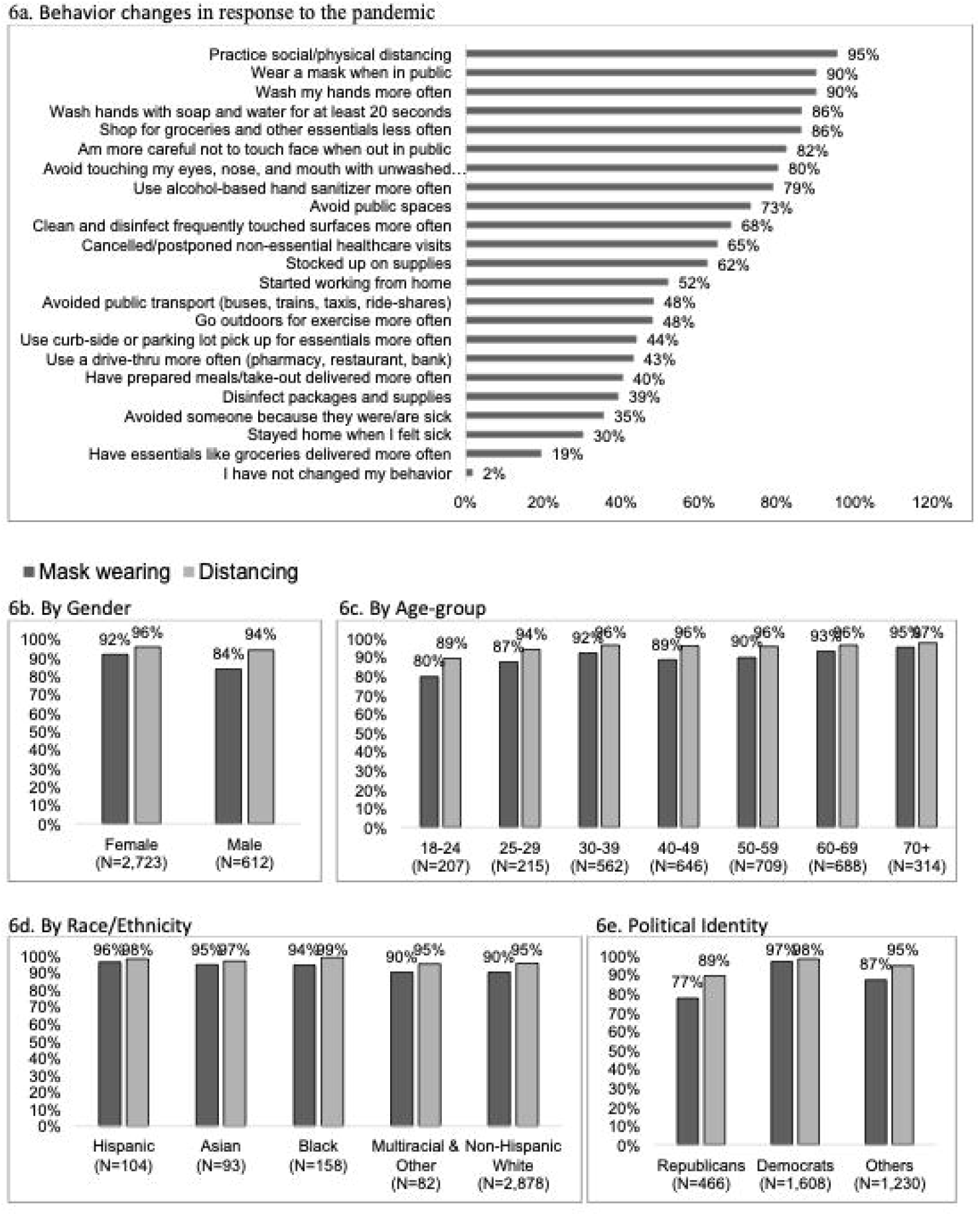
Behavior changes in response to the pandemic and masking and distancing in public by participant characteristics. Survey responses to the question: “How (if at all) have you changed your behavior in response to the coronavirus/COVID-19? (Check all that apply)” for all respondents (6a). Percent of respondents reporting mask wearing and distancing by gender (6b), age-group (6c), race/ethnicity (6d), political identity (6e), education level (6f), income level (6g), and Virginia region (6h). Percent of respondents reporting an information source as trustworthy by if they reported wearing or not wearing a mask in public (6i) and by distancing or not distancing in public (6j).

Wearing a mask in public was reported by more women than men (92% vs. 84%) (see Fig 6b), and increased by age from 80% of those 18-24 years old to 95% of those 70 years and older (see Fig 6c), more Democrats than Republicans and others (97%, 77%, and 87%, respectively), increased by education level from 76% of those with a high-school education or less to 94% of those with a doctoral degree, and more higher- than middle- and lower-income (92%, 89%, 87%). Those in Southwest Virginia reported less mask wearers (86%) than other regions (91%-95%) (see Fig 6f). Distancing was more common than masking in all groups, but showed similar demographic trends as wearing a mask.

More of those who reported wearing vs. not wearing a mask reporting national health and science organizations (89% vs. 58%), state or local health departments (78% vs. 51%), health care professional (75% vs 64%), online news (57% vs. 38%), local government leaders (49% vs. 24%), local TV news (35% vs. 26%), local newspaper (21% vs. 6%), and radio (21% vs. 13%) as a trusted source of information (see Fig 6i). A smaller proportion of those who reported wearing vs. not wearing a mask reported the federal government as a trusted source (21% vs. 28%) or not following COVID-19 information (1% vs. 12%). Similar trends were observed for distancing (see Fig 6j).

In adjusted logistic analyses, we found that the odds of reporting not wearing a mask in public was greater than their comparative groups for those living in Southwest Virginia vs. all other regions combined (OR= 1.95, 95% CI= 1.48, 2.58), men vs. women (OR= 1.85, 95% CI= 1.36, 2.51), young adults vs. other age-groups combined (OR= 2.42, 95% CI= 1.54 vs. 3.81), non-Hispanic Whites vs. other races combined (odds ratio [OR]=1.68, 95% CI= 1.04, 2.71), and those with household income under $100,000 vs. those with income at least $100,000 (OR= 1.41; 95% CI= 1.06, 1.89) (see Table 2). The odds of reporting not wearing a mask in public was greater for those identifying as a Republican vs. Democrat (OR=5.42, 95% CI= 3.63, 8.09), those who did not vs. did report national science and health organization(s) as a trusted information source (OR= 3.16, 95% CI= 2.21, 4.51), those who believed one or more alternative messages vs. not believing in any (OR= 2.09, 95% CI= 1.48, 2.94). Not having a college degree was associated with not wearing a mask in unadjusted analyses (OR= 1.98, 95% CI= 1.54, 2.55) but in adjusted analyses (OR= 0.98, 95% CI= 0.71, 1.35). All other associations were statistically significant (p<0.05) in both adjusted and unadjusted analyses. Odds ratios for distancing showed similar associations as for masking, but at a smaller magnitude. Region, gender, race/ethnicity, and education were not statistically significant (p<0.05) in adjusted analyses for distancing.

**Table 2.** Odds ratios (ORs) of reporting not wearing masks in public and not practicing social/physical distancing using logistic regression with robust standard errors (N=3,307).

| Variables | Unadjusted<br>OR (95% CI); p-value | Adjusted*<br>OR (95% CI); p-value | Unadjusted<br>OR (95% CI);<br>p-value | Adjusted*<br>OR (95% CI); p-value |
| --- | --- | --- | --- | --- |
| Southwest vs. all other regions | 2.33 (1.84, 2.96);<br>**<0.001** | 1.95 (1.48, 2.58);<br>**<0.001** | 1.44 (1.02, 2.03); **0.037** | 1.13 (0.75, 1.70);<br>0.567 |
| Men vs. women | 2.14 (1.66, 2.76);<br>**<0.001** | 1.85 (1.36, 2.51);<br>**<0.001** | 1.48 (1.01, 2.18); **0.046** | 1.32 (0.82, 2.15);<br>0.255 |
| 18-24 vs. all other age-groups | 2.59 (1.90, 3.71);<br>**<0.001** | 2.42 (1.54, 3.81);<br>**<0.001** | 2.89 (1.79, 4.66); **<0.001** | 2.34 (1.34, 4.11);<br>**0.003** |
| Non-Hispanic White vs. other races | 1.63 (1.09, 2.46);<br>**0.018** | 1.68 (1.04, 2.71);<br>**0.035** | 1.66 (0.91, 3.01); 0.099 | 1.83 (0.92, 3.63);<br>0.083 |
| No college degree vs. college degree | 1.98 (1.54, 2.55);<br>**<0.001** | 0.98 (0.71, 1.35);<br>0.897 | 3.52 (2.52, 4.92); **<0.001** | 1.46 (0.96, 2.25);<br>0.080 |
| Less than \$100,000 vs. \$100,000 or more | 1.67 (1.31, 2.13);<br>**<0.001** | 1.41 (1.06, 1.89);<br>**0.020** | 2.11 (1.47, 3.03); **<0.001** | 1.62 (1.03, 2.54);<br>**0.035** |

| Political Identity |  |  |  |  |
| --- | --- | --- | --- | --- |
| Democrat | Ref | Ref | Ref | Ref |
| Republican | 8.79 (6.18, 12.48);<br>**<0.001** | 5.42 (3.63, 8.09);<br>**<0.001** | 6.92 (4.31, 11.1); **<0.001** | 3.53 (2.06, 6.07);<br>**<0.001** |
| Independent, other, no preference | 4.44 (3.21, 6.13);<br>**<0.001** | 3.16 (2.21, 4.51);<br>**<0.001** | 3.15 (2.01, 4.93); **<0.001** | 1.79 (1.07, 3.01);<br>**0.026** |
| Not reporting national science and health organizations as trusted source of information | 5.21 (4.14, 6.54);<br>**<0.001** | 3.45 (2.57, 4.66);<br>**<0.001** | 5.57 (4.20, 7.41); **<0.001** | 2.96 (1.95, 4.49);<br>**<0.001** |
| Belief in alternative message(s) | 3.39 (2.65, 4.33);<br>**<0.001** | 2.09 (1.48, 2.94);<br>**<0.001** | 4.17 (3.09, 5.64); **<0.001** | 2.65 (1.66, 4.23);<br>**<0.001** |

\*Adjusted models include all variables listed in the table (N=2,992 with complete information for all variables). P-values <0.05 are in bold.

## Discussion

In a convenience sample of adults residing in Virginia, we found many differences in where people received information that they trust, what they believed, and how their behaviors changed in response to the COVID-19 pandemic by socio-demographics, political identity, and geography within Virginia. Respondents who identified as non-Hispanic White, men, Republican, other political identity, younger age, income <$100,000, did not report national science and health organizations as a trusted source, reported believing an alternative message, and/or living in Southwest Virginia had greater odds of not wearing a mask than their comparative groups in both unadjusted and adjusted logistic regression. Differences in physical difference were also observed for physical distancing for these same variables, but at a lower magnitude as distancing was more likely than masking across all groups so differences were less pronounced.

Our study was subject to several limitations. First, we conducted numerous comparisons and did not adjust for multiple comparisons due to the exploratory nature of this study and some statistically significant associations could be due to chance. Second, complete demographic and socioeconomic information was missing from 9% of respondents included in this study. Third, the political identity response options were limited to Republican, Democrat, independent, and other, resulting in individuals identifying as “independent” and “other” being grouped together, although these individuals may hold extremely diverse political views. Finally, while we were able to make comparisons between subgroups, our internet-based convenience sample is not representative of the generalized Virginia population (United States Census Bureau 2019) and may not reflect conditions at other time points given that the survey was conducted in the summer of 2020. Data collection began on May 19^th^, just prior to the racial justice protests that began on May 26^th^ in Minneapolis and continued throughout the United States [37]. People’s behavior may have been altered based on their cost-benefit analyses of COVID-19 risk and the risks associated with racial injustice over the course of our data collection period [38, 39].

Other cross-sectional survey studies from early in the COVID-19 pandemic (spring to summer 2020) produced similar results [10, 29-36]. For example, studies in Australia, Malaysia, Italy, Canada, the United Kingdom, Arab countries, and other regions of the United States found that evidence-based COVID-19 messaging significantly impacted respondents’ beliefs and risk mitigation behaviors. Multiple studies showed that consistent messaging focusing on positive ways to cope with lockdowns and other COVID-19 mitigation measures were more effective than messaging focused only on compliance in promoting long-term behavioral changes like staying at home, mask-wearing, social distancing, and hand-washing [29-31, 36]. Studies also showed that while older adults were generally more concerned and had higher anxiety levels about potential COVID-19 infection, they were also less concerned than younger adults about the short- and long-term economic instabilities caused by the pandemic [30-33, 36, 40]. Other studies also found that women, racial/ethnic minorities, and those with lower socioeconomic status experienced more COVID-19 anxieties compared to men, ethnic majorities, and those of higher socioeconomic status [10, 33, 34, 36]. In multiple countries, people identifying as politically conservative and those with lower health literacy and education level were more likely to report not following recommended COVID-19 precautions and believing that people were overreacting [29, 32-34, 41]. People identifying as politically liberal and those with higher health literacy and education were more likely to follow public health guidelines and believe that their governments were not doing enough to stop the pandemic [32-34, 36]. Multiple studies showed that misinformation exposure and beliefs were consistently higher among younger people, ethnic minorities, and those who identified as politically conservative [33, 34, 41-43]. Several preliminary studies have also shown that misinformation and mistrust in government entities and/or the vaccine development process are major contributing factors to vaccine hesitancy, especially among minority populations and people with low education levels, socioeconomic status, and low perceived risk of contracting COVID-[44-46]. Our study supports and adds to this knowledge describing how information sources considered trustworthy vary across these different population impacting believes and behaviors.

This study can assist decision makers and the public in developing more effective public health messaging for both the ongoing COVID-19 pandemic and for future public health challenges in Virginia and similar settings in the United States. Future studies could include quantitative subgroup, subregion, and qualitative analyses to enhance our understanding of the nuances related to designing effective public health messaging.

## Data Availability

Data cannot be shared publicly because of confidentiality of sensitive data. Data are available by contacting the corresponding authors and completing a data sharing agreement.

## Acknowledgements

R Silverman assisted in the study design and implementation, performed the statistical analysis, interpreted the data, and wrote the paper. D Short assisted in the study implementation and helped write the paper. S Wenzel assisted in the study design and implementation and helped write the paper. M Friesen assisted in the study design and implementation, and helped write the paper. N Cook is the principal investigator for this project, assisted in the study design and implementation, and helped write the paper. We thank the other members of our study team: Kristin Miller, Emily Wells, Kathy Hosig, Amy Smith, Amanda Nguyen, Molly Roberts, Teace Markwalter, and Kristina Jiles for their assistance with survey distribution and recruitment. Thank you to Luela Ba and Becky Willis for helping make tables and figures. We thank the survey respondents for their participation.

