## supplement for "COVID-19 related messaging, beliefs, information sources, and mitigation behaviors in Virginia: A cross-sectional survey in the summer of 2020"

### COVID-19 Messaging Survey

---

**Start of Block: Default Question Block**

Q1 A team of researchers in Virginia would like to learn about how Virginians heard and responded to information about the **coronavirus**/**COVID-19**. We will use the results from this survey to help inform and improve public health messages in Virginia.

Your participation in this Virginia Tech Research Survey on Covid-19/Coronavirus Messaging is voluntary and involves no more than minimal risk. All responses are **anonymous** and **confidential** and cannot be traced back to you. This survey should take about **5 to 10 minutes** to complete. You may skip any questions you feel uncomfortable answering. We may present or publish the results so that they may benefit other communities. If you have questions about your rights as a research participant, or concerns or complaints about the research, you may contact the Virginia Tech HRPP at 540-231-3732 or and reference the IRB number #20-353
OR
Project Investigator: Dr. Natalie E. Cook, Do you consent (agree) to participate in this anonymous survey?

○ No (4)

○ Yes (5)

*Skip To: End of Survey If A team of researchers in Virginia would like to learn about how Virginians heard and responded to... = No*

*Skip To: Q45 If A team of researchers in Virginia would like to learn about how Virginians heard and responded to... = Yes*

---

Q45 Are you 18 years or older?

○ No (1)

○ Yes (4)

*Skip To: End of Survey If Are you 18 years or older? = No*

*Skip To: Q47 If Are you 18 years or older? = Yes*

Q47 Do you currently live in Virginia?

- ◯ No (1)
- ◯ Yes (3)

*Skip To: End of Survey If Do you currently live in Virginia? = No*

*Skip To: Q28 If Do you currently live in Virginia? = Yes*

Q28 The following messages are related to the coronavirus/COVID-19 (***not all are true***). Please check all that apply if you have **heard, believe,** and/or **changed your behavior** based on each message.

*If you are using a phone to complete this survey, please click each message below to expand and check the options that fit for you.*

| | I have **heard** this message (1) | I **believe** this message (2) | This message has **affected what I do** (4) |
|---|---|---|---|
| The coronavirus is highly contagious (1) | ☐ | ☐ | ☐ |
| Flatten the curve (2) | ☐ | ☐ | ☐ |
| Stay home and stay safe (3) | ☐ | ☐ | ☐ |
| COVID-19 was developed to increase sales of cleaning supplies or medicine (4) | ☐ | ☐ | ☐ |
| There is no cure for COVID-19 (5) | ☐ | ☐ | ☐ |
| Stay home, save lives, slow the spread (6) | ☐ | ☐ | ☐ |
| Practice social distancing (7) | ☐ | ☐ | ☐ |
| Don't touch your face (9) | ☐ | ☐ | ☐ |
| Your mask protects me and my mask protects you (10) | ☐ | ☐ | ☐ |
| COVID-19 was developed for population control (11) | ☐ | ☐ | ☐ |
| Wash your hands for at least 20 seconds (12) | ☐ | ☐ | ☐ |
| COVID-19 can be treated with natural remedies, herbs, or teas (13) | ☐ | ☐ | ☐ |

| | | | |
|---|---|---|---|
| COVID-19 is a hoax (14) | ☐ | ☐ | ☐ |
| The coronavirus/COVID-19 is an apocalyptic/end-of-times sign (15) | ☐ | ☐ | ☐ |
| The coronavirus was developed as a bioweapon (16) | ☐ | ☐ | ☐ |
| COVID-19 was developed to lower social security payments to senior citizens (17) | ☐ | ☐ | ☐ |
| The coronavirus/COVID-19 is just like the flu (18) | ☐ | ☐ | ☐ |

Q3 Where do you get information that you trust about **coronavirus/COVID-19**? (Check all that apply)

- ☐ Family/friends  (1)
- ☐ Healthcare professional  (2)
- ☐ Faith leader (such as pastor, clergy, imam, priest, rabbi, etc.)  (3)
- ☐ Local news on TV  (16)
- ☐ National news on TV  (17)
- ☐ Local printed newspaper  (19)
- ☐ Radio  (20)
- ☐ Online news  (21)
- ☐ Social media (such as Facebook, Twitter, etc.)  (22)
- ☐ Local government leaders  (23)
- ☐ Federal government leaders (such as the White House)  (24)
- ☐ National science/health organizations (such as Dr. Fauci from the National Institutes of Health (NIH), Centers for Disease Control (CDC), etc.)  (25)
- ☐ State or local health department  (26)
- ☐ I do not follow coronavirus/COVID-19 updates  (27)
- ☐ Other (please specify)  (28) ________________________________________________

*Display This Question:*

*If Where do you get information that you trust about coronavirus/COVID-19? (Check all that apply) = Local news on TV*

Q25 Which **local** TV news channel(s) do you watch for information that you trust about coronavirus/COVID-19? (Check all that apply)

- ☐ ABC (1)
- ☐ CBS (2)
- ☐ FOX (3)
- ☐ NBC (6)
- ☐ PBS (8)
- ☐ Other (please specify) (5) ________________________________________________

---

*Display This Question:*

*If Where do you get information that you trust about coronavirus/COVID-19? (Check all that apply) = National news on TV*

Q48 Which **national** TV news channel(s) do you watch for information that you trust about coronavirus/COVID-19?
(Check all that apply)

- ▢ ABC (1)
- ▢ CBS (2)
- ▢ FOX News (8)
- ▢ CNN (3)
- ▢ PBS (9)
- ▢ MSNBC (7)
- ▢ FOX (4)
- ▢ NBC (6)
- ▢ Other (please specify) (5) ________________________________________________

---

*Display This Question:*

*If Where do you get information that you trust about coronavirus/COVID-19? (Check all that apply) = Radio*

Q26 Which radio station(s) do you listen to for information that you trust about coronavirus/COVID-19?

________________________________________________________________

---

*Display This Question:*

*If Where do you get information that you trust about coronavirus/COVID-19? (Check all that apply) = Online news*

Q27 Which online news site(s) do you read for information that you trust about coronavirus/COVID-19?

________________________________________________________________

*Display This Question:*

*If Where do you get information that you trust about coronavirus/COVID-19? (Check all that apply) = Social media (such as Facebook, Twitter, etc.)*

Q25 Which social media platform(s) do you use for information that you trust about coronavirus/COVID-19?
(Check all that apply)

▢ Facebook (1)

▢ Twitter (2)

▢ Other (please specify) (3) ________________________________________________

Q40 How (if at all) have you changed your behavior in response to the **coronavirus/COVID-19**?
(Check all that apply)

- ▢ Started working from home (2)
- ▢ Shop for groceries and other essentials less often (3)
- ▢ Stocked up on supplies (4)
- ▢ Practice social/physical distancing (stay at least 6 feet away from others, except people I live with) (5)
- ▢ Wash my hands with soap and water for at least 20 seconds (6)
- ▢ Use alcohol-based hand sanitizer more often (20)
- ▢ Avoid touching my eyes, nose, and mouth with unwashed hands (19)
- ▢ Wash my hands more often (18)
- ▢ Clean and disinfect frequently touched surfaces more often (22)
- ▢ Am more careful not to touch face when out in public (7)
- ▢ Have prepared meals/take-out delivered more often (8)
- ▢ Have essentials like groceries delivered more often (9)
- ▢ Use curb-side or parking lot pick up for essentials more often (15)
- ▢ Use a drive-thru more often (such as at the pharmacy, restaurant, or bank) (10)
- ▢ Disinfect packages and supplies (21)
- ▢ Go outdoors for exercise more often (11)

☐ Wear a mask when in public (14)

☐ Avoid public spaces (16)

☐ Cancelled/postponed non-essential healthcare visits (17)

☐ Stayed home when I felt sick (23)

☐ Avoided someone because they were/are sick (24)

☐ Avoided public transportation such as buses, trains, taxis, or ride-shares (25)

☐ I have not changed my behavior (1)

☐ Other (please specify) (13) ________________________________________________

---

Q52 In the past 2 weeks (14 days), how many times did you **go into** a retail location, such as a grocery store or drug store?
*Do not include curbside pick-up of items purchased without entering the store.*

○ none in the past 2 weeks (1)

○ 1 time (2)

○ 2 times (3)

○ 3 times (4)

○ 4 times (5)

○ 5 times (6)

○ 6 or more times (7)

---

Q30 How serious do you think the **coronavirus/COVID-19** is?

- ○ Not at all serious (1)
- ○ Somewhat serious (13)
- ○ Very serious (14)
- ○ I'm not sure (15)

---

Q4 Which (if any) of the following have affected whether or not you think the **coronavirus/COVID-19** is serious?
(Check all that apply)

- ☐ I became sick (1)
- ☐ Someone I knew became sick (2)
- ☐ I am or live with someone who is high-risk (such as being elderly or having a chronic condition) (17)
- ☐ Public schools closed (3)
- ☐ Restaurant dining rooms shut down (4)
- ☐ VA Governor Northam declared a state of emergency (5)
- ☐ I began to work from home (6)
- ☐ I was furloughed (laid off) from work or lost my job (7)
- ☐ The CDC recommended that everyone wear face masks (8)
- ☐ Hearing about COVID-19 in other countries (9)
- ☐ Hearing about COVID-19 in other states (10)
- ☐ Stores began limiting purchase amounts of household items (11)
- ☐ Church/religious gatherings were moved online (12)
- ☐ Sporting events were cancelled or postponed (NBA, MLB, March Madness, the Olympics, etc.) (18)
- ☐ A celebrity, famous athlete, politician, or other public figure tested positive for COVID-19 (19)

- ☐ VA Governor Northam issued a **recommendation** that Virginians stay at home (13)
- ☐ VA Governor Northam issued an official **mandated** stay-at-home order (14)
- ☐ I do not think coronavirus/COVID-19 is serious (20)
- ☐ Other (please specify) (15) ________________________________________________

---

Q54 How worried are you about catching the coronavirus/COVID-19?

- ○ Not at all worried (2)
- ○ Somewhat worried (3)
- ○ Very worried (1)

---

Q56 How worried are you about severe disease or complications if you were to catch the coronavirus/COVID-19?

- ○ Not at all worried (1)
- ○ Somewhat worried (2)
- ○ Very worried (3)

---

Q53 Have you been tested for the coronavirus/COVID-19 by a medical doctor?

- ○ **Yes**, I **was tested**, and the test was **positive** for COVID-19 (1)
- ○ **Yes**, I **was tested**, and the test was **negative** for COVID-19 (2)
- ○ **Yes**, I **was tested**, and I **do not want to share** the results (5)
- ○ **No**, I tried to get tested but **could not** get a test (3)
- ○ **No**, I **have not tried** to get tested (4)

---

Page Break

Q7 Almost done! Please answer the following questions about yourself. Remember that this survey is anonymous, so we will not be able to trace your answers back to you.

---

*

Q8 What is your zip code?

________________________________________________________________

---

Q11 What is your gender?

- ◯ Female (1)
- ◯ Male (2)
- ◯ Prefer to self-describe (please specify) (3) ________________________________________________

---

Q43 What is your age group?

- ◯ 18-24 (1)
- ◯ 25-29 (2)
- ◯ 30-39 (3)
- ◯ 40-49 (4)
- ◯ 50-59 (5)
- ◯ 60-69 (10)
- ◯ 70-79 (11)
- ◯ 80+ (12)

---

Q29 What is the highest level of school you have completed or the highest degree you have received?

- Less than high school degree (1)
- High school diploma or GED (2)
- Some college but no degree (3)
- Trade school or Associate degree (4)
- Bachelor's degree (5)
- Master's degree (6)
- Doctoral or professional degree (7)

Q37 What categories best describes you?
(Check all that apply)

- ☐ American Indian or Alaska Native (1)
- ☐ Asian (2)
- ☐ Black or African-American (3)
- ☐ Hispanic, Latinx, or Spanish origin (4)
- ☐ Middle Eastern or North African (5)
- ☐ Native Hawaiian or Other Pacific Islander (6)
- ☐ White (7)
- ☐ Other race, ethnicity, or origin (please specify) (8) ________________________________________________

---

Q21 Generally speaking, do you usually think of yourself as a Republican, a Democrat, an Independent, or something else?

- ○ Republican (1)
- ○ Democrat (2)
- ○ Independent (3)
- ○ Other (please specify) (4) ________________________________________________
- ○ No preference (5)

---

Q51 Which of the following best describes you?

- ◯ Heterosexual or straight (1)
- ◯ Homosexual, gay, or lesbian (2)
- ◯ Bisexual (3)
- ◯ I'm not sure (5)
- ◯ Prefer to self-describe (please specify) (4) ________________________________________________

---

Q17 What was your estimated household income last year (before taxes)?

- ◯ Less than $20,000 (1)
- ◯ $20,000 to $39,999 (2)
- ◯ $40,000 to $59,999 (3)
- ◯ $60,000 to $79,999 (7)
- ◯ $80,000 to $99,999 (9)
- ◯ $100,000 or more (11)

---

Q19 What is your current employment status?

☐ Full-time (1)

☐ Part-time (2)

☐ Seeking opportunities (3)

☐ Retired (5)

☐ Not working due to disability (6)

☐ Student (12)

☐ Not employed (please specify reason) (14) ________________________________________________

☐ Other (please specify) (7) ________________________________________________

Q35 How has your employment status or income changed due to the **coronavirus/COVID-19**?

○ No change in employment/income (1)

○ I permanently lost my primary source of employment/income (2)

○ I temporarily lost my primary source of employment/income (such as being furloughed) (3)

○ My employment/income was reduced (6)

○ I gained employment/income (4)

○ Other (please specify) (5) ________________________________________________

Q50 Which best describes your primary occupation or job sector?

*If retired or recently unemployed, please indicate your occupation/job sector before*

*unemployment.*
*(Choose one)*

- ○ Administrative and support services (37)
- ○ Agriculture, forestry, fishing and hunting (24)
- ○ Accommodation (41)
- ○ Arts, entertainment and recreation (40)
- ○ Childcare provider (8)
- ○ Construction (26)
- ○ Custodian (21)
- ○ Day-laborer (19)
- ○ Educational services (38)
- ○ Facilities (50)
- ○ Finance and insurance (33)
- ○ Food service (47)
- ○ Gig work/freelance (12)
- ○ Grocery/food retail (5)
- ○ Health care (provider) (39)
- ○ Health care (office and administrative) (53)
- ○ Information (32)
- ○ Management of companies and enterprises (36)
- ○ Manufacturing (27)
- ○ Mining quarrying, and oil and gas extraction (25)

- ◯ Military (44)
- ◯ Office and administrative support (other than heath care) (16)
- ◯ Public services or administration (42)
- ◯ Personal care and services (15)
- ◯ Professional, scientific, and technical services (35)
- ◯ Public health (52)
- ◯ Real estate and rental and leasing (34)
- ◯ Retail trade (other than grocery/food) (29)
- ◯ Social services (49)
- ◯ Trade (such as electrician, plumber, etc.) (23)
- ◯ Transportation of materials and warehousing (20)
- ◯ Transportation of people (51)
- ◯ Utilities (31)
- ◯ Waste management services (48)
- ◯ Wholesale trade (28)
- ◯ Other (please specify) (46) ________________________________________________

---

Q38 Is your current job classified as "essential" during the COVID-19 pandemic?

- ◯ Yes (1)
- ◯ No (2)
- ◯ I don't know (3)

Q21 Which of the following best describes your relationship status?

- ○ Single (1)
- ○ Partnered, not living together (2)
- ○ Partnered, living together (3)
- ○ Married (4)
- ○ Separated or divorced (5)
- ○ Widowed (6)

Q27 Including yourself, how many people live in your household?

- ○ 1 (4)
- ○ 2 (5)
- ○ 3 (6)
- ○ 4 (7)
- ○ 5 (8)
- ○ 6 (9)
- ○ 7 (10)
- ○ 8 (11)
- ○ 9 (12)
- ○ 10 or more (13)

Q27 Are you or someone in your household at high-risk (being elderly, having a chronic condition such diabetes, heart disease, high blood pressure, or lung problems, immunosuppressed, or pregnant)?

- Yes (1)
- No (2)
- I'm not sure (4)

---

Q49 Do you currently live with or care for any children (under 18 years old)?

- Yes (1)
- No (4)

---

Page Break

Q6 Do you have anything else to share about how you heard and responded to information about the coronavirus/COVID-19?

________________________________________________________________

________________________________________________________________

________________________________________________________________

________________________________________________________________

________________________________________________________________

**End of Block: Default Question Block**
